# Multimodal FDG-PET and EEG assessment improves diagnosis and prognostication of disorders of consciousness

**DOI:** 10.1101/2020.06.16.20132514

**Authors:** Bertrand Hermann, Johan Stender, Marie-Odile Habert, Aurélie Kas, Mélanie Denis-Valente, Federico Raimondo, Pauline Pérez, Benjamin Rohaut, Jacobo Diego Sitt, Lionel Naccache

## Abstract

The complex diagnosis of disorders of consciousness (DoC) diagnosis increasingly relies on brain-imaging techniques for their ability to detect residual signs of consciousness in otherwise unresponsive patients. However, few of these techniques have been validated on external datasets. Here, we show that the FDG-PET glucose metabolic index of the best preserved hemisphere has robust in-sample and out-sample performances to diagnose DoC, slightly outperforming EEG-based classification. We further show that a multimodal assessment combining both FDG-PET and EEG not only improved diagnostic performances, but also allowed to identify covert cognition and to predict 6-month responsiveness in initially unresponsive patients. Lastly, we show that DoC heterogeneity reflects a sum of regional cortical metabolic differences, and their corresponding behavioral patterns, rather than a binary contrast between conscious and unconscious states. In total, we show that FDG-PET and EEG provide complementary information on DoC physiopathology and that their combination improves DoC diagnosis and prognostication.

## INTRODUCTION

Disorders of consciousness (DoC) are common consequences of severe brain injury. They comprise heterogeneous conditions, with patients either suffering from a complete loss of awareness of themselves and their environment, as in the vegetative state (also termed the unresponsive wakefulness syndrome – VS/UWS) (Jennett and Plum, 1972; Laureys *et al*., 2010), or patients exhibiting minimal but definite signs of external awareness, as in the minimally conscious state (MCS) (Giacino *et al*., 2002). The differential diagnosis is mainly based on systematic behavioral assessments, among which the coma recovery scale – revised (CRS-R) is the current gold-standard (Kalmar and Giacino, 2005). This scale scores behavior in response to hierarchically ordered prompts in six domains: auditory, visual, motor, oromotor and verbal, communication and arousal.

We recently proposed a reinterpretation of the MCS concept, on the basis that the clinical signs defining the condition do not necessarily reflect minimal awareness, but rather index the presence of cortically-driven behavior. MCS should therefore be viewed as a cortically-mediated state (CMS) (Naccache, 2018) rather than a specific state of consciousness. In keeping with this view, recent advances indicate that purely behavioral evaluation of consciousness may be inadequate, as residual signs of awareness can be identified using brain-imaging techniques in ~15-25% of clinically unresponsive patients (Owen *et al*., 2006; Cruse *et al*., 2012; King *et al*., 2013). The presence of covert cognition furthermore appears to hold important prognostic significance (Sitt *et al*., 2014; Claassen *et al*., 2019). In light of both these considerations, neuroimaging tools are essential both to complement the clinical diagnosis and to investigate the neurophysiological basis of the disorders (Hermann *et al*., 2020). Among the available tools, 18F-fluoro-deoxyglucose positron emission tomography (FDG-PET) have consistently demonstrated a metabolic reduction to approximately 50% of brain glucose uptake across various unconscious conditions, from anesthesia, to sleep, to DoC. As such it has been proposed as a diagnostic tool for DoC patients (Stender *et al*., 2014, 2015). However, in the aforementioned studies, diagnostic labels relied on the subjective assessment of PET results and/or seed-based normalization procedures, prone to odd results in this specific population. In 2016, Stender *et al*., proposed a novel normalization procedure resulting in a simple measure of cerebral metabolic activity, which showed the best diagnostic performances to date (Stender *et al*., 2016). Yet, this measure has never been validated outside the initial center. Since the normalization procedure requires on-site acquisition of healthy controls, its generalizability is still unknown.

Apart from FDG-PET, other diagnostic tools, which are easier to implement at bedside, have been proposed. Electrophysiology allows reliable and robust automatic classification of conscious versus unconscious states based both on EEG brain activity (Sitt *et al*., 2014; Engemann *et al*., 2018) and event related potentials (ERP) during cognitive tasks such as the auditory local-global paradigm (Bekinschtein *et al*., 2009; Faugeras *et al*., 2011, 2012). As these techniques develop and become more readily available, it is increasingly necessary to understand the interrelation of the physiological data they provide. In this context, glucose metabolism as measured by FDG-PET, has been previously associated with EEG-derived network metrics of interconnected central hubs (Chennu *et al*., 2017) and correlated with resting-state fMRI connectivity maps (Soddu *et al*., 2016). While such comparative studies remain rare, they are nonetheless critical to the wider implementation of neuroimaging- and electrophysiological techniques to routine clinical use.

We here aimed to determine the clinical utility of the previously proposed FDG-PET metabolic index for DoC diagnosis. We performed an external validation of the PET metabolic index to diagnose MCS, and subsequently compared its in-sample and out-sample performances to EEG-based classification. We further explored whether a multimodal approach combining PET and EEG could outperform any of these two single measures. Finally, in light of our proposed development of the CMS concept, we investigated the regional metabolic patterns associated with the MCS condition, to better understand the neuro-metabolic difference between MCS and VS/UWS.

## MATERIAL AND METHODS

### Ethics statement

The protocol conformed to the Declaration of Helsinki, to the French regulations, and was approved by the local ethic committee (*Comité de Protection des Personnes n*° *2013-A01385-40*) Ile de France 1 (Paris, France) under the code *‘Recherche en soins courants’* (routine care research). Informed consent was obtained from each patient’s relative.

### Population

We prospectively included patients with disorders of consciousness, admitted to the Neurology Intensive Care Unit of the Pitié-Salpêtrière university hospital (Paris, France). All patients were transferred for specialized diagnosis, in order to determine their state of consciousness. The admission lasted approximately one week, during which they were subject to repeated behavioral assessment, a high-density EEG recording and an FDG-PET. Previously acquired PET images from 32 healthy subjects with no history of neurological disorders were used as controls.

### Behavioral assessment

We assessed the DoC patients’ behavior using the gold-standard JFK Coma Recovery Scale-Revised (CRS-R)(Giacino and Kalmar, 2005). This scale evaluates the patient’s response to a set of hierarchically ordered items in six different domains: auditory, visual, motor, language and oromotor, communication and arousal. During the hospitalization, several CRS-R were performed by trained physicians in order to increase diagnostic accuracy. The reference standard was the highest detected level of consciousness, as defined by the best response obtained among all CRS-R scorings. We collected the 6-month outcome by phone interview of the treating physician and/or family. Since recovery of consciousness was a rare event in such a short-term outcome for patients suffering from chronic disorders of consciousness, we focused on a more reasonable yet clinically relevant outcome, namely: the recovery of responsiveness in initially unresponsive patients (responsiveness was defined as the reproducible response to command following the scoring guidelines of the CRS-R).

### PET acquisition and image analysis

#### Acquisition

All PET images were acquired on a Philips Gemini GXL scanner (Philips Medical Systems) in the nuclear medicine department of the hospital. Patients were without mechanical ventilation and unsedated for at least 48h prior to acquisition. Patients and controls received a bolus injection of FDG adjusted to body weight (2 MBq/kg) and were kept at rest in a dark and quiet room. PET images were recorded starting from 20 minutes to 1 hour after the circulation of the tracer using two different protocols: the first consisted in the static acquisition of a single 15 minutes frame, the second, used in restless patients in order to minimize motion artifacts, consisted in the acquisition of three consecutives frames of 5 minutes each. Images were then reconstructed using iterative LOR-RAMLA algorithm (2 iterations), with a « standard » post-reconstruction filter. All corrections (attenuation, scatter and random coincidence) were integrated in the reconstruction.

#### Quantitative normalization procedure

The quality of PET images was assessed by nuclear medicine physicians, blinded to the patients state of consciousness. PET data of insufficient quality for the standard visual interpretation were discarded. PET acquired more than one hour after the tracer injection were also discarded according to standard clinical practice. The uptake quantification procedure followed steps described by Stender *et al*. (Stender *et al*., 2016). The image is hereby normalized to match uptake-histograms of extracerebral cephalic tissues, as opposed to common methods normalizing brain metabolism to a specific cerebral region (such as the cerebellum). First, images were registered to a common template in MNI space by affine and non-linear transformation. They were then segmented (left and right cerebral cortices and extracerebral tissue) and normalized on the metabolism of the extracerebral tissue in reference to controls (by minimization of the Jensen-Shannon divergence between patients and the control distribution). Finally, brain metabolic activity was scaled by setting the mean activity of extracerebral regions to an index value of one. Metabolic index of the best preserved hemisphere (MIBH) was computed as the highest mean metabolic activity of the two hemispheres. For the dynamic acquisition, the quantification procedure was performed on each one of the three frames and the resulting normalized images were then averaged.

### EEG

#### Acquisition and preprocessing

High-density scalp EEG sampled at 250 Hz were recorded at bedside the during the ‘local-global’ paradigm (see below) using a Net300 Amplifier and 256 electrodes HydroCel Geodesic Sensor Net (Electrical Geodesics, Eugene, Oregon, USA). Impedances were set below 100 kΩ prior to acquisition. EEG were preprocessed using a previously described pipeline (Engemann *et al*., 2015, 2018), with the segmentation of the recordings in epochs according to the onset of the fifth sound (from 800ms before to 740 ms after) followed by an automated rejection of artifacted epochs and/or channels procedure based on maximal amplitude (150 µV) and variance (z-score=4) of the EEG signal. Recordings with more than 30% of channels and/or 70% of epochs rejected were discarded and bad channels of the remaining recordings were interpolated.

#### Auditory oddball local and global effect

The local-global paradigm is an auditory oddball paradigm proposed to probe the unconscious and conscious processing of auditory novelty, through the manipulation of two temporal levels of auditory regularities violation, respectively on a short-time scale (local) or on a long-time scale (global). The latter is able to elicit the P3b signature of conscious auditory processing (Bekinschtein *et al*., 2009). Epochs were baseline corrected over the 800 ms window preceding the onset of the fifth sound and t-tests for unequal variance were conducted over the whole time-series for both local and global contrasts. To determine the presence of a local and global effect in individual subjects, we used the previously published stringent triple-threshold criteria (Faugeras *et al*., 2012). An effect was considered present, if a significant difference (p-value ≤ 0.01) was observed on at least 5 consecutive time samples and 10 adjacent electrodes starting from the onset of the fifth sound and if this difference was stronger than any differences observed during the baseline period (either lower minimal p-value or if equal, longer duration). Accordingly, three groups of patients were identified: absent local and global effect, local effect only and global effect (regardless of the presence of a local effect, as the global effect indexes a higher level in the auditory novelty processing hierarchy).

#### Automatic classification of conscious states

112 features reflecting averages and fluctuations in time and space of 28 EEG markers of spectral power, connectivity, complexity and evoked responses were computed from the 800 ms baseline period of the local-global EEG recordings (Sitt *et al*., 2014; Engemann *et al*., 2018). These features were then used to predict the patients state of consciousness using a linear support vector machine (SVM) algorithm as previously published(Sitt *et al*., 2014).

### Statistical analysis

#### Diagnostic performances

We assessed in-sample and out-of-sample diagnostic performances of two index tests, the FDG-PET MIBH and the EEG-based prediction of consciousness, using the standard following discrimination metrics with their 10000 bootstrapped 95% confidence interval (CI95%): area under the ROC curves (AUC), sensitivity, specificity, positive and negative predictive values, positive and negative likelihood ratios, and accuracy. For the FDG-PET, in-sample performances were derived from the optimal MIBH threshold according to the ROC curve, while out of sample performances were derived from the use of the 3.18 MIBH threshold value set by Stender *et al*. (Stender *et al*., 2016). For in-sample EEG performances, the SVM algorithm was trained and tested on the same patients, namely the cohort of patients included in the study. Out of sample performances were obtained by training the SVM classifier on a previously published dataset from which we excluded the patients included in the study to avoid overfitting, yielding a total of 341 recordings (VS/UWS n=170, MCS n=171 acquired from 267 independent subjects (Engemann *et al*., 2018). Predicted probability of MCS classification (as opposed to VS/UWS) was obtained from the SVM output through Platt scaling. Since this output is probabilistic, discrimination metrics were computed for a 0.5 threshold in both conditions. ROC curves comparisons were tested using 10000 bootstrapped replicates. Sensitivities and specificities were compared using McNemar test for paired design. Quantitative data were expressed as mean ± standard deviation or median [interquartile range] and compared through Student t-test and Mann-Whitney-U and categorical data were expressed as number (percentage) and compared through chi-squared test or Fisher’s exact test as appropriate. Correlation between the MIBH and the CRS-R was performed using spearman correlation coefficient.

#### PET regional analysis

In order to investigate whether a classification based on regional metabolism would outperform the MIBH (which measures the hemispheric cortical average), we extracted the average metabolic index values for each patient within each of the 41 cortical regions of each hemisphere, as defined by the Automated Anatomical Labeling (AAL) atlas in MNI152 space (Tzourio-Mazoyer *et al*., 2002). For each of these regions we then computed the AUC of VS/UWS vs. MCS discrimination with two methods (bootstrapped CI95% and permutation testing against 0.5 with 10000 replicates).

#### PET voxel-based analysis

To study the regional metabolic patterns associated with the MCS-specific behavioral signs, we then performed a voxel-wise analysis of the metabolic index using a linear model. Each subscale of the CRS-R was dichotomized according to the presence (1) or absence (0) of MCS items (except for the attentional subscale which was dichotomized according to: <2=0, ≥2=1), and these scores were included in the model as independent variables. The independent effect of the presence of MCS items in each subscale was then sequentially tested on the whole brain with the metabolic index of each voxel as the dependent variable. For these analyses, images were smoothed with an 8mm full-width at half-maximum gaussian kernel, and masked using a gray matter MNI template. Significance threshold was set to p<0.005, uncorrected, with a minimum extent of 100 voxels per cluster.

#### Softwares

The PET quantification procedure used the following softwares: MRIcron software package (MRIcron version 6 june 2013, McCausland Center, University of South Carolina), Advanced Normalization Tools (ANTs version 2.0.3), Matlab (Matlab 8.4 Mathworks Inc., Natick, Massachusetts) with data from the SPM8 Matlab toolbox (Statistical Parametric Mapping version 8, Wellcome Trust Centre for Neuroimaging, University of London). EEG and regional and voxel-wise PET analyses were done in Python (version 3.6.7) using the free *NICE library* (Engemann *et al*., 2018), https://github.com/nice-tools/nice, *MNE-python* (Gramfort *et al*., 2013), *scikit-learn* (Pedregosa *et al*., 2011), *nilearn* (Abraham *et al*., 2014) and *nistats* packages. Diagnostic performances were computed in R (version 3.3.2 (2016-10-31) with *caret, DTComPair, pROC* (Robin *et al*., 2011) and *epiR* packages.

### Data and software availability

Data supporting the findings of the study are available from the corresponding author, upon reasonable request.

## RESULTS

Between January 2016 and October 2019, we evaluated 182 independent DoC patients, of which 89 were eligible for PET (no mechanical ventilation and no sedation in the preceding 48h) and 74 were actually scanned (see flow chart Figure 1). Five recordings were discarded because they were scanned more than one hour after the tracer injection, four were discarded due to poor image quality, and one recording was not included because the patient was comatose at the time of scanning. The final dataset comprised FDG-PET recordings of 64 individual DoC patients: 23 in VS/UWS, 34 in MCS and 7 in EMCS. Diagnostic performance measures of the FDG-PET and EEG were restricted to the VS/UWS and MCS population. As expected, significant differences existed between VS/UWS and MCS patients in terms of etiology (p <10^−3^), with more anoxia in VS/UWS (65%) than in MCS (17%), while injury of vascular origin showed the reverse pattern (0% vs. 24%) and CRS-R scores (6 [5-7] vs. 11 [9-13] for VS/UWS and MCS respectively, p<10^−3^). Importantly, no significant differences were found between VS/UWS and MCS patients in terms of number of CRS-R performed, CRS-R arousal score, delay between PET and best CRS-R and PET acquisition parameters (acquisition protocol, tracer dose and time from tracer injection), except for the blood glucose concentration which remained within normal range (5.9 [5.2-6.6] in VS/UWS patients vs. 5.3 [4.8-5.8] in MCS patients, p=0.019, Table 1) and which did not impact classification performances (see below).

**Table 1.**
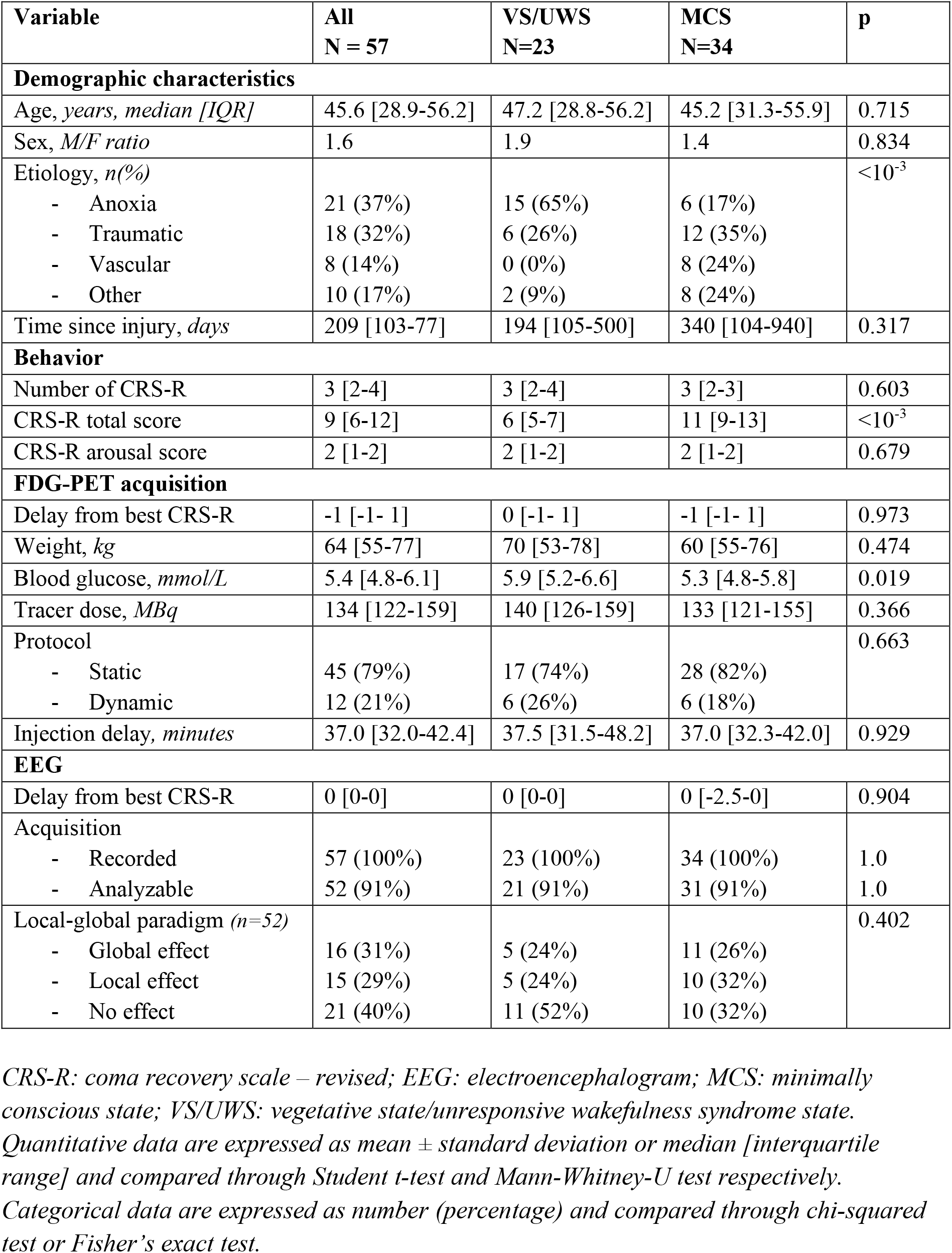
FDG-PET Population characteristics

**Figure 1.**
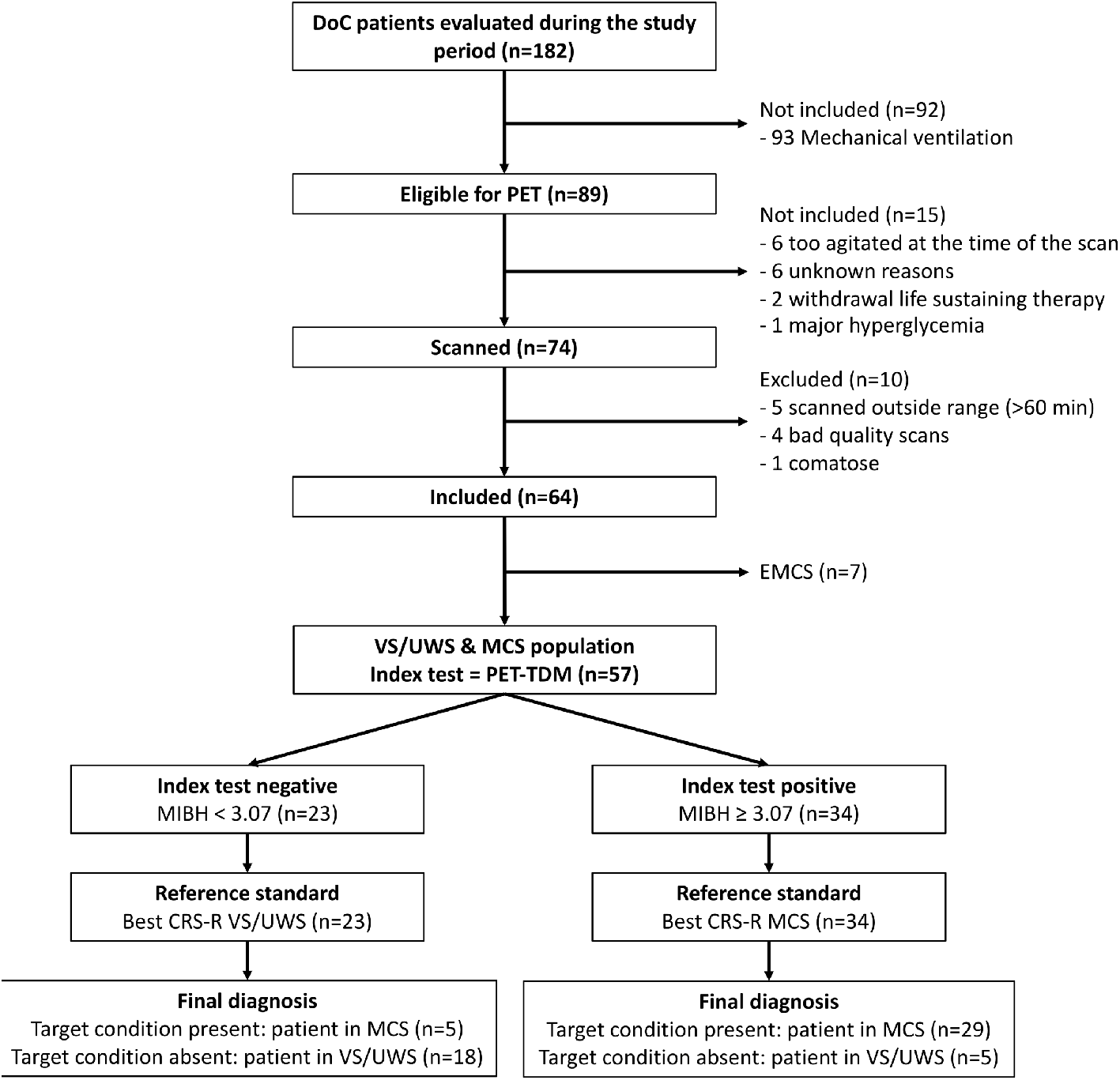
Flow chart. CRS-R: coma recovery scale – revised; EMCS: emergence of minimally conscious state; MCS: minimally conscious state; MIBH: metabolic index of the best preserved hemisphere.

### Validation of the FDG-PET metabolic index as a reliable diagnostic tool

We first evaluated the in-sample FDG-PET performances of the MIBH to diagnose MCS, as compared to the reference gold standard, i.e. the best state of consciousness observed over a range of repeated CRS-R measurements. VS/UWS patients had a significantly lower MIBH than MCS patients (median MIBH of 2.70 vs. 3.65, p<10^−4^), with a good discrimination performances (AUC 0.821 [0.694-0.930]). At the optimal MIBH cut-off of 3.07, corresponding to 54% of the healthy controls metabolism (median MIBH of 5.73), accuracy was 84% [71-92], positive predictive value was 85% [69-95], negative predictive value was 78% [56-93], sensitivity was 85% [69-95] and specificity was 78% [56-93]. Similar discrimination performances were found when including blood glucose as covariate in the model and when adjusting for the scanning protocol (see Supplementary Material).

External validation of the PET using the previously published diagnostic threshold, based on data from Liège University Hospital (MIBH=3.18) still showed good discrimination performances with an accuracy of 79% [66-89], positive predictive value of 84% [67-95], negative predictive value of 72% [51-88], sensitivity of 79% [62-91] and specificity 78% [56-93]. As expected, all EMCS patients had a score above threshold (mean MIBH of 4.39) and interestingly, all MCS patients with a score below threshold were MCS *minus* patients (MCS-), that is MCS patients without clinical evidence of language preservation, contrary to MCS *plus* patients (MCS+) (Figure 2A). Finally, in addition to its classification performance, MIBH also correlated strongly with the CRS-R score (spearman ρ=0.59, p<10^−4^, Figure 2B). These reliable performances validate the FDG-PET as a robust method to diagnose MCS across centers.

**Figure 2.**
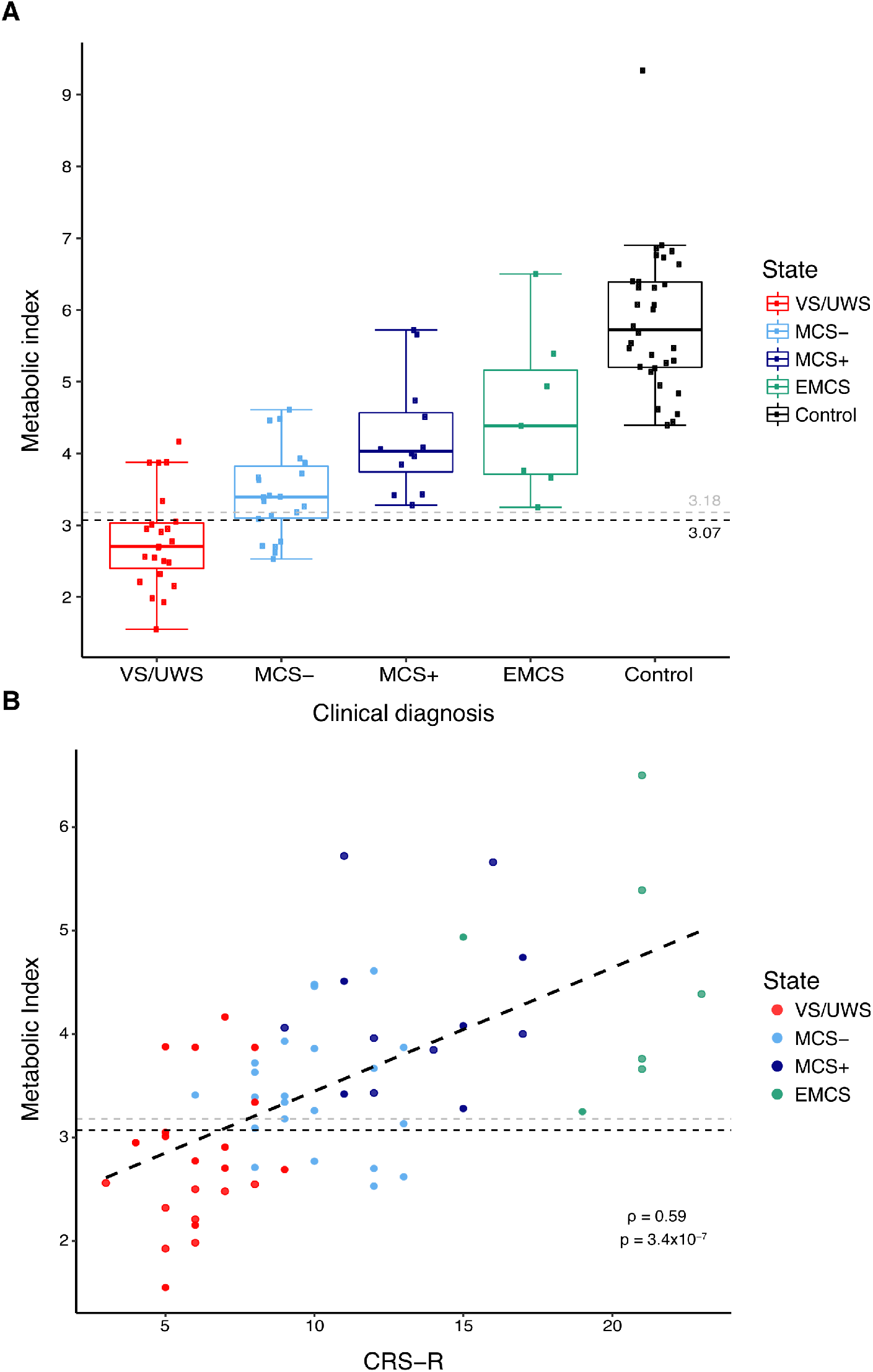
FDG-PET metabolic index and consciousness. **(A)** Higher metabolic index of the best preserved hemisphere values are observed with increasing state of consciousness, from VS/UWS, MCS-, MCS+ to EMCS patients (control patients used for the normalization are in black). Dashed lines optimal in-sample (black) and out-of-sample (gray) thresholds for VS/UWS discrimination. **(B)** Significant correlation of the metabolic index with the CRS-R scores, Spearman’s rho=0.59, p<10^−4^.

### FDG-PET slightly outperforms EEG-based classification

In order to evaluate the added value of the FDG-PET, we compared its diagnostic performances to a validated and robust EEG-based classification tool (Sitt *et al*., 2014) which demonstrated its ability to generalize to different settings: centers, paradigms, number of EEG channels and length of recordings (Engemann *et al*., 2018). Among the 57 EEG recordings, 5 did not pass the automated preprocessing and quality control pipeline, leaving a population of 52 patients (21 VS/UWS and 31 MCS) in which we compared FDG-PET and EEG performances (Table 2 and Figure 3). In-sample FDG-PET performances in this population were similar to the one of the whole PET population and EEG and FDG-PET performances were not significantly different (p=0.218). EEG out-of-sample discrimination performances were: AUC of 0.770 [0.619-0.896], accuracy of 67% [53-80], sensitivity of 58% [39-75] and specificity of 81% [58-95]. While AUC were not significantly different (p=0.628), the FDG-PET had a higher sensitivity than EEG (paired McNemar test p=0.033) with no difference in specificity (paired McNemar test p=0.655).

**Table 2.** Diagnostic performances of PET-FDG and EEG

| IN-SAMPLE PERFORMANCES |  |  |  |  |  |  |  |  |  |  |  |  |
| --- | --- | --- | --- | --- | --- | --- | --- | --- | --- | --- | --- | --- |
| Imaging Metrics | FDG-PET |  |  |  | EEG |  |  |  | FDG-PET \| EEG |  |  |  |
| Contingency table |  | MCS | VS | All |  | MCS | VS | All |  | MCS | VS | All |
|  | + | 27 | 5 | 32 | + | 29 | 6 | 35 | + | 31 | 8 | 39 |
|  | - | 4 | 16 | 20 | - | 2 | 15 | 17 | - | 0 | 13 | 13 |
|  | All | 31 | 19 | 52 | All | 31 | 21 | 52 | All | 31 | 21 | 52 |
| AUC | 0.816 [0.681-0.928] |  |  |  | 0.912 [0.807-0.986] |  |  |  | 0.810 [0.714-0.905] |  |  |  |
| Accuracy | 83 [70-92] |  |  |  | 85 [72-93] |  |  |  | 85 [72-93] |  |  |  |
| Sensitivity | 87 [70-96] |  |  |  | 94 [79-99] |  |  |  | 100 [89-100] |  |  |  |
| Specificity | 76 [53-92] |  |  |  | 71 [48-89] |  |  |  | 62 [38-82] |  |  |  |
| Positive PV | 84 [67-95] |  |  |  | 83 [66-93] |  |  |  | 79 [64-91] |  |  |  |
| Negative PV | 80 [56-94] |  |  |  | 88 [64-99] |  |  |  | 100 [75-100] |  |  |  |
| Positive LR | 3.66 [1.68-7.96] |  |  |  | 3.27 [1.65-6.48] |  |  |  | 2.62 [1.52-4.53] |  |  |  |
| Negative LR | 0.17 [0.07-0.44] |  |  |  | 0.09 [0.02-0.35] |  |  |  | 0.00 [0.00-Inf] |  |  |  |
| OUT-OF-SAMPLE PERFORMANCES |  |  |  |  |  |  |  |  |  |  |  |  |
| Imaging Metrics | FDG-PET |  |  |  | EEG |  |  |  | FDG-PET \| EEG |  |  |  |
| Contingency table |  | MCS | VS | All |  | MCS | VS | All |  | MCS | VS | All |
|  | + | 26 | 5 | 31 | + | 18 | 4 | 22 | + | 29 | 7 | 36 |
|  | - | 5 | 16 | 21 | - | 13 | 17 | 30 | - | 2 | 14 | 16 |
|  | All | 31 | 21 | 52 | All | 31 | 21 | 52 | All | 31 | 21 | 52 |
| AUC | 0.816 [0.681-0.928] |  |  |  | 0.770 [0.619-0.896] |  |  |  | 0.801 [0.689-0.905] |  |  |  |
| Accuracy | 82 [69-91] |  |  |  | 67 [53-80] |  |  |  | 83 [70-92] |  |  |  |
| Sensitivity | 84 [66-95] |  |  |  | 58 [39-75] |  |  |  | 94 [79-99] |  |  |  |
| Specificity | 76 [53-92] |  |  |  | 81 [58-95] |  |  |  | 67 [43-85] |  |  |  |
| Positive PV | 84 [66-95] |  |  |  | 82 [60-95] |  |  |  | 81 [62-99] |  |  |  |
| Negative PV | 76 [53-92] |  |  |  | 57 [37-75] |  |  |  | 88 [62-98] |  |  |  |
| Positive LR | 3.52 [1.61-7.69] |  |  |  | 3.05 [1.20-7.73] |  |  |  | 2.81 [1.52-5.17] |  |  |  |
| Negative LR | 0.21 [0.09-0.49] |  |  |  | 0.52 [0.33-0.82] |  |  |  | 0.10 [0.02-0.38] |  |  |  |

*EEG: electroencephalogram; LR: likelihood ratio; MCS: minimally conscious state; PV: predictive value; VS: vegetative state/unresponsive wakefulness syndrome state; |: and/or; +: test positive; - : test negative.*
*Values are expressed with their 95% confidence interval.*

**Figure 3.**
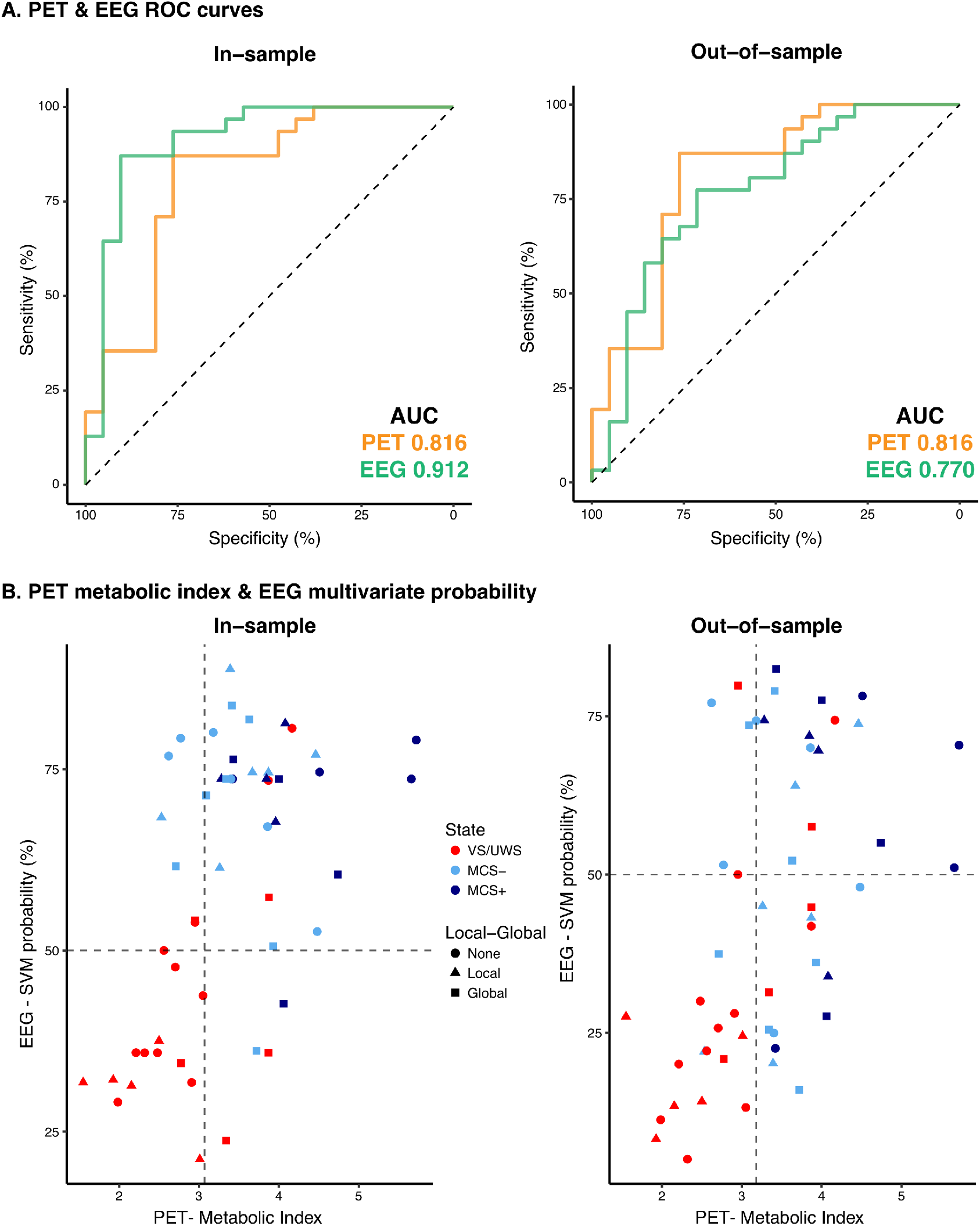
FDG-PET and EEG performances. ROC curves of in-sample (left) and out-of-sample (right) diagnostic performances of FDG-PET (yellow) and EEG-based classification (green) with corresponding discrimination area under the curve (AUC). Similar in-sample AUC were found for FDG-PET and EEG (0.816 vs. 0.912, p=0.218) while FDG-PET slightly outperformed EEG, however not significantly, on out-sample performances (0.816 vs. 0.770, p=0.628). **(B)** Scatterplot representation of the support vector machine (SVM) classifier probability to be classified MCS according to the FDG-PET metabolic index of the best preserved hemisphere (MIBH) and the absence (circle) or presence of unconscious (local effect, triangle) or conscious (global effect, square) auditory processing of auditory novelty during the ‘local-global’ paradigm. Dashed gray lines represent thresholds for MCS vs. VS/UWS discrimination.

### Combination of FDG-PET and EEG identifies covert consciousness and improves prognostication

Ideally, a perfect brain-imaging diagnostic tool for DoC patients should first identify all clinically MCS patients. We thus tested if multimodal brain-imaging combining FDG-PET and EEG (MIBH higher than Liège threshold for MCS and/or EEG-based predicted probability of MCS >0.5) could outperform any of the two imaging modality taken in isolation.

The combination of the PET metabolic index with the EEG-based classification indeed accurately identified almost all MCS patients, yielding an improved sensitivity of 94% [79-99%] with a specificity of 67% [43-85], a positive predictive value of 81% [62-99] and negative predictive value of 88% [62-98]. Sensitivity of the combination was significantly higher than the one of EEG alone (paired McNemar test p<0.001) and there was a trend towards a higher sensitivity than FDG-PET alone (paired McNemar test p=0.083). But more importantly, since potential cognitive-motor dissociation has been demonstrated (Owen *et al*., 2006; Cruse *et al*., 2012; Schiff, 2015; Edlow, 2018; Claassen *et al*., 2019), the specificity of brain-imaging techniques in comparison of the CRS-R gold-standard may not be an adequate metric to measure the ability of a new diagnostic tool to detect residual signs of cognition. We thus investigated the potential added value of the combination of FDG-PET and EEG in unresponsive patients using two measures independent from the CRS-R and relevant to the diagnosis of consciousness: an ERP neural correlate of conscious auditory perception and 6-month outcome. In this context, 7 out of the 21 (33%) clinically VS/UWS patients exhibited a higher metabolism and/or richer brain electrophysiological activity than expected. Interestingly 4 of these (57%) also exhibited an ERP global effect that indicates preserved conscious processing of auditory novelty (Bekinschtein *et al*., 2009; Faugeras *et al*., 2011). This was in contrast to the very low proportion of patients classified as VS/UWS by both behavior and neuro-imaging techniques and showing a global effect (1 out of the 14 (7%), fisher exact test p=0.025). Moreover, the combination of PET and EEG was significantly associated with the recovery of responsiveness at 6 months in initially unresponsive patients (9/24 (38%) of patients with high FDG-PET metabolism and/or rich EEG activity had recovered responsiveness at 6 months vs. 0/16 (0%) of patients with low-level FDG-PET metabolism and EEG activity, fisher exact test p=0.006), while either FDG-PET or EEG alone were not significantly associated with 6-month responsiveness (7/19 (37%) vs. 2/21 (10%), fisher exact test p=0.060 for FDG-PET and 5/13 (38%) vs. 4/27 (15%), fisher exact test p=0.120 for EEG). These results strongly suggest that the combination of FDG-PET and EEG accurately identified residual signs of high cognitive function fostering the recovery of responsiveness in otherwise clinically unresponsive patients.

### Significance of the VS/UWS – MCS distinction

#### Regional metabolism discrimination performances

We subsequently investigated whether any regional brain metabolism region could outperform the MIBH. To this end, we computed the AUC from average metabolic values of 41 cortical regions in both hemispheres. We found that all cortical regions significantly discriminated VS/UWS from MCS patients (all false-discovery rate corrected p-values <0.05), and that several regions had similar (or even slightly better) performances than the MIBH (Figure 4, Supplementary Table 2). Importantly, the latter included primary or secondary sensory areas, not specifically associated with consciousness: the left paracentral lobule (AUC 0.835 [0.730-0.919]), the left lingual and calcarine regions from the occipital cortex (AUCs 0.834 [0.731-0.918] and 0.832 [0.728-0.922] respectively) as well as the left and right supplementary motor areas (AUCs 0.817 [0.699-0.911] and 0.816 [0.701-0.909] respectively). Actually, among the regions traditionally associated with consciousness, only the left precuneus ranked in the top 10 discriminative regions (AUC 0.821 [0.705-0.912]). These findings suggest that the VS/UWS vs. MCS contrast reflect an aggregate of cortical network activity, across a multitude of brain functions, rather than a pure minimal contrast between a conscious and unconscious state.

**Figure 4.**
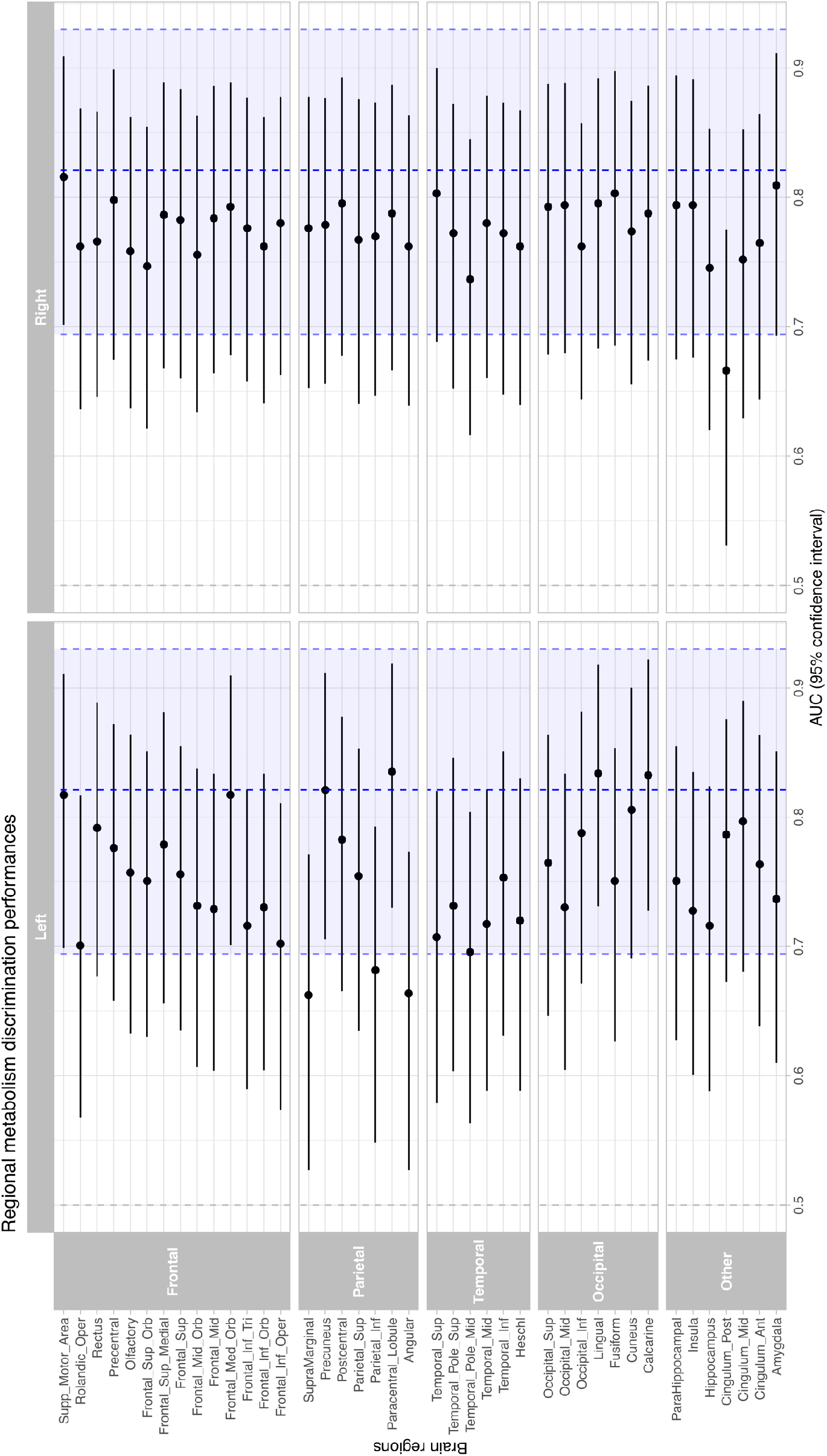
FDG-PET regional discrimination performances. Respective AUCs and 10000 bootstrapped 95% confidence intervals for the VS/UWS vs. MCS discrimination of the 41 cortical regions of the AAL atlas in both hemispheres.10000 permutation testing against 0.5, all false discovery rate (FDR) corrected p-values < 0.05. Blue dashed line and shaded region represent the AUC and 95% confidence interval of the metabolic index of the best preserved hemisphere.

#### MCS items are associated with metabolic specific of subscales

Following this line of reasoning, we investigated the metabolic correlates of the CRS-R, which is the gold-standard to define the states of consciousness. We dichotomized each CRS-R subscale according to the presence or absence of an MCS item in each individual, and used parametric statistical mapping to investigate the specific metabolic pattern associated with each subscale. This analysis showed that the presence of a visual MCS item or a motor MCS item were significantly associated with metabolism restricted in first-order cortical areas, occipital cortex and motor and premotor cortices respectively (Figure 5), without activation in associative prefrontal or parietal cortices. On the contrary, the presence of the auditory response to command MCS item (either reproducible or systematic), was significantly associated with a higher metabolism in widespread cortical areas, encompassing left-lateralized temporal and frontal language related regions, temporal pole as well as the dorsolateral prefrontal cortex and posterior cingulate/precuneus. Notably no significant differences were observed for either the communication or oromotor/verbal function subscale, potentially due to the rarity of patients exhibiting these behaviors (N=5 and N=1 respectively). These distinct patterns of metabolic activity consistent with the observed behaviors, further stress the heterogeneity of MCS patients and the difficult interpretation of CRS-R items in terms of consciousness physiology.

**Figure 5.**
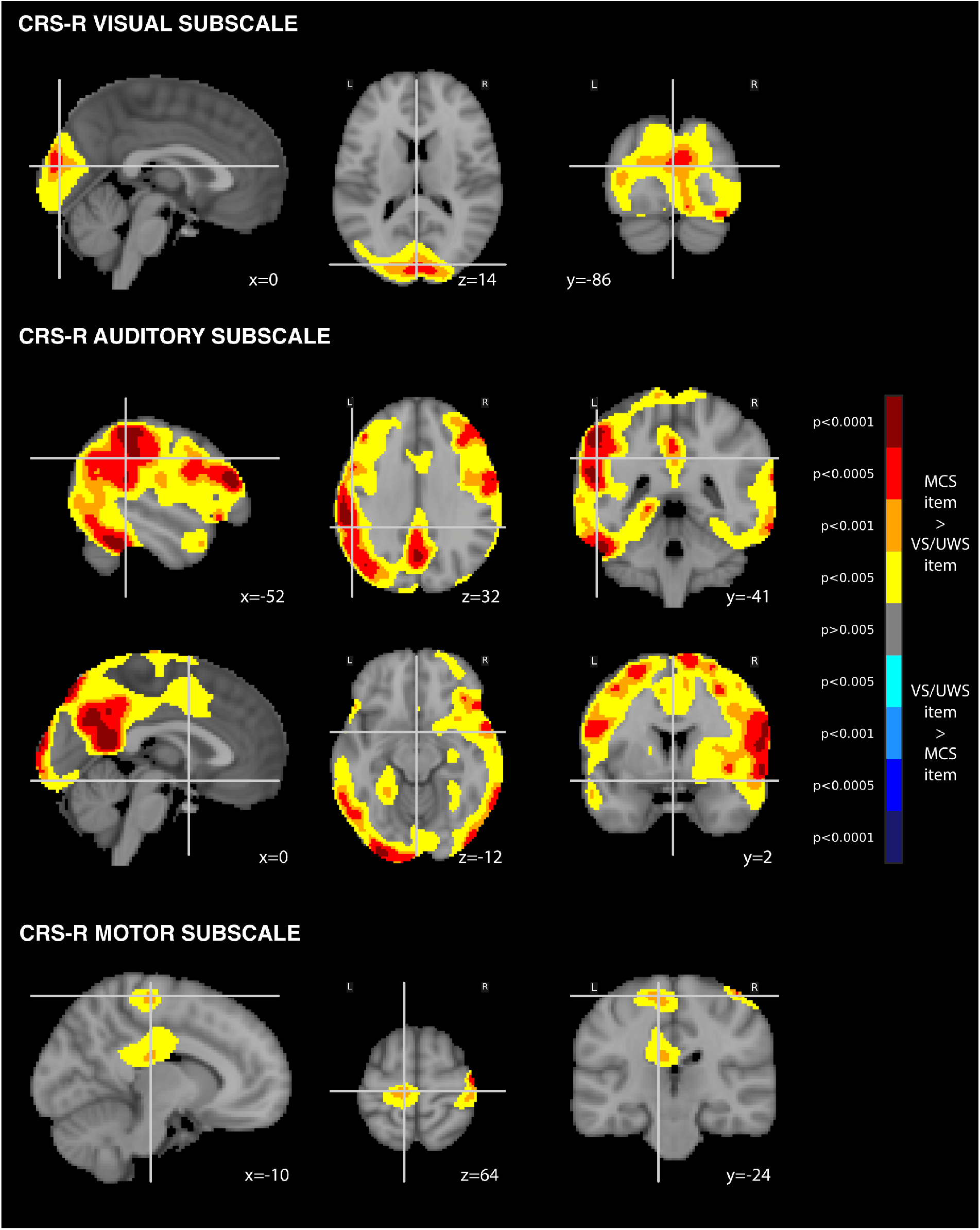
Metabolic correlates of CRS-R MCS items. Independent FDG-PET metabolic correlates of the CRS-R MCS items in **(A)** the auditory subscale **(B)** the visual subscale and **(C)** the motor subscale (p<0.005 uncorrected, cluster extent 100 voxels, superimposed on coronal, sagittal and axial slices of the MNI 152 T1 brain template with related y, x and z MNI coordinates). L=left; R=right.

## DISCUSSION

### FDG-PET metabolic index is an accurate and robust procedure to diagnose MCS

In this study, we showed that external implementation and generalization of the metabolic index quantification procedure of FDG-PET glucose uptake was practically feasible, and enabled accurate diagnosis of disorders of consciousness. In our cohort, we found that the approximate metabolic boundary between MCS and VS/UWS was 50% of normal metabolism, slightly higher than previously found by Stender *et al*. (Stender *et al*., 2016). While this difference could be due to site and/or protocol specific factors, our results align well with previous findings in DoC patients, sleep and anesthesia (Braun *et al*., 1997; Maquet *et al*., 1997; Nofzinger *et al*., 2002; Laureys *et al*., 2004; Shulman *et al*., 2009; Laureys and Schiff, 2012). More generally, these differences could be due to the natural variability of brain metabolism even among normal healthy subjects. All in all, our results confirm that quantitative FDG-PET using the metabolic index quantification procedure is easy to apply, accurate and robust across sites to diagnose the state of consciousness in this severely brain-injured population.

### Multimodal brain-imaging improves diagnosis and prognostication of disorders of consciousness

We first showed that FDG-PET performed slightly better than EEG to diagnose DoC, with a difference in performance seemingly driven by a greater number of EEG-based false-negatives; clinically MCS patients estimated by the EEG classification to be unconscious. This phenomenon may be explained by common fluctuations of awareness among the MCS patients (Wannez *et al*., 2017). While FDG-PET integrates brain activity across tens of minutes and could therefore be less susceptible to these fast and transient changes, EEG records brain activity in the ms range and is likely more affected. Yet, EEG availability and robust performances (Engemann *et al*., 2018) still make it a great candidate to bedside diagnostic assessment of patients, at least as a first-line/screening diagnostic procedure. Indeed, FDG-PET logistic requirements exceed those of bedside EEG, and it should be noted that even in our expert center, PET-scanning of mechanically ventilated patients was not possible. Besides, whereas FDG-PET provides information on the localization and amount of neuronal firing, it does not inherently provide insights into the underlying information processing. In contrast, EEG explores the qualitative features of the information exchange, but has limited anatomical resolution and the signal is difficult to quantify. Thus EEG and FDG-PET theoretically capture partially overlapping, yet independent, features of the DoC pathophysiology, which could complement each other. Although the good diagnostic agreement between the two modalities fulfills the need of consilience between brain-imaging procedures to address the diagnostic challenge of cognitive motor dissociation (Peterson, 2016), the combination of both techniques clearly improves both the diagnosis and the prognostication of disorders of consciousness. First, this combination satisfies the criterion of maximization of MCS diagnosis sensitivity, but most importantly, it allows to identify covert cognition in otherwise unresponsive patients. Although, our study does not provide definite answers as to the cognitive status of the seven clinically VS/UWS patients, who were classified as MCS using both PET and EEG, it is notable that their proportion corresponds to previous reports on the prevalence of covert cognition after severe brain injury (Kondziella *et al*., 2016). Besides, more than half of these patients with absent behavioral responsiveness but rich PET/EEG activity showed an ERP neural signature of conscious access in the local-global paradigm. In contrast, only 1 of the 14 behaviorally VS/UWS patients confirmed by PET/EEG showed this global effect signature (Bekinschtein *et al*., 2009; Faugeras *et al*., 2011, 2012; Raimondo *et al*., 2017), suggesting a covert cognitive state richer than the one inferred from their behavior, ie. a state of cognitive-motor and higher-order motor dissociations (Schiff, 2015; Edlow *et al*., 2017). This is further supported by the higher recovery of responsiveness at 6 months observed in initially unresponsive patients with rich PET and/or EEG activity. This finding is in line with previous studies showing that both FDG-PET (Stender *et al*., 2014, 2016) and EEG-based classification (Sitt *et al*., 2014; Chennu *et al*., 2017; Claassen *et al*., 2019) convey meaningful prognostic information. Here, again, we show that the combination of PET and EEG outperform both modalities in isolation. Overall, our results provide evidence in favor of the multimodal assessment of disorders of consciousness, as previously proposed theoretically (Bayne *et al*., 2017; Naccache, 2018). Such assessment should combine different clinical and brain-imaging techniques to investigate the degree of preservation of neuronal architecture involved in conscious processing and consciousness recovery.

### FDG-PET supports the concept of cortically-mediated state

One of the major advantage of the FDG-PET over EEG relates to its high spatial resolution enabling analyses of regional metabolic patterns. Using a cortical atlas, we showed that the distinction of MCS from VS/UWS patients could be accurately achieved from each separate cortical region. While the left precuneus, - a region crucial to conscious processing (Boly *et al*., 2008; Vanhaudenhuyse *et al*., 2010; Crone *et al*., 2015) - exhibited high discrimination performance, this was also the case for primary and secondary specialized cortical areas such as occipital or supplementary motor areas. These results are in accordance with a previous report of the VS/UWS vs. MCS contrast (Stender *et al*., 2014). Moreover they complement the previous demonstration that several fMRI resting state networks, including auditory, sensorimotor and visual networks were also accurate in discriminating MCS from VS/UWS (Demertzi *et al*., 2015). This spurred us to investigate the regional cortical patterns independently associated with MCS items in each CRS-R subscale. This analysis showed that the most prevalent MCS items, namely MCS visual and to a lesser extent motor items (Wannez *et al*., 2018) were associated with metabolic activity within specialized and restricted occipital and frontal regions, that do not correspond to the fronto-parietal network typically observed during conscious states (Maquet *et al*., 1997; Nofzinger *et al*., 2002; Laureys *et al*., 2004; Boveroux *et al*., 2010; Laureys and Schiff, 2012). By contrast, auditory subscale response to command, close to the reportability criteria definition of consciousness, was associated not only with metabolic activity in language related regions, but also in the default-mode network, which is closely related to conscious processing. This shows that MCS behavioral heterogeneity is paralleled by a corresponding cortical metabolism heterogeneity, supporting the cortically-mediated state hypothesis. In this view, the current complex of behaviors defining MCS is not specific to the residual consciousness of patients, but rather inform us with some confidence about the functional preservation of certain cortical networks (Naccache, 2018). While a seemingly trivial insight at first glance, the modern brain imaging techniques have already sparked a reappraisal of a clinical semiology developed decades ago. The physiological significance of visual fixation is for example a point of current debate. Indeed, the absence of metabolic differences between MCS patients with fixation only and VS/UWS patients cast doubt on the relevance of this sign as an MCS item (Bruno *et al*., 2010). This physiological heterogeneity may also explain the inter-subject variability in response to certain treatments. As such, recent studies demonstrated a correlation between the therapeutic efficacy of transcranial direct current stimulation, and the subjects underlying brain anatomy and metabolism (Thibaut *et al*., 2015, 2018; Hermann *et al*., 2019). More developed neuroimaging techniques could both help to better stratify patients for therapeutic intervention, and to develop tailored treatment strategies, which are still desperately needed for these debilitating conditions.

## CONCLUSION

FDG-PET pseudo-quantitative metabolic index of the best preserved hemisphere is an accurate and robust procedure across sites to diagnose MCS, which can even be improved in combination with EEG-based classification allowing the detection of covert cognition and 6-month responsiveness recovery in unresponsive patients. Moreover, our results strongly suggest that the behavioral diagnosis of MCS does not correspond to an elusive and generic conscious state, but rather to a CMS that reveals the preservation of metabolic activity in specialized cortical networks. In total, we show that FDG-PET and EEG provide complementary information on DoC physiopathology that may be combined to improve their diagnosis and prognostication.

## Data Availability

Data supporting the findings of the study are available from the corresponding author upon reasonable request.

## Declaration of interest

None declared

## SUPPLEMENTARY MATERIAL

### Supplementary results

1. **EEG/PET population characteristics (Supplementary Table 1)**
2. **Influence of blood glucose on FDG-PET diagnostic performances**
3. **PET regional metabolism discrimination performances (Supplementary Table 2)**
4. **Metabolic correlates of CRS-R motor and arousal MCS items with liberal statistical threshold (Supplementary Figure 1)**
5. **Whole-brain voxel-wise state of consciousness comparisons (Supplementary Figure 2)**

### 1. EEG/PET population characteristics (Supplementary Table 1)

**Supplementary Table 1.**
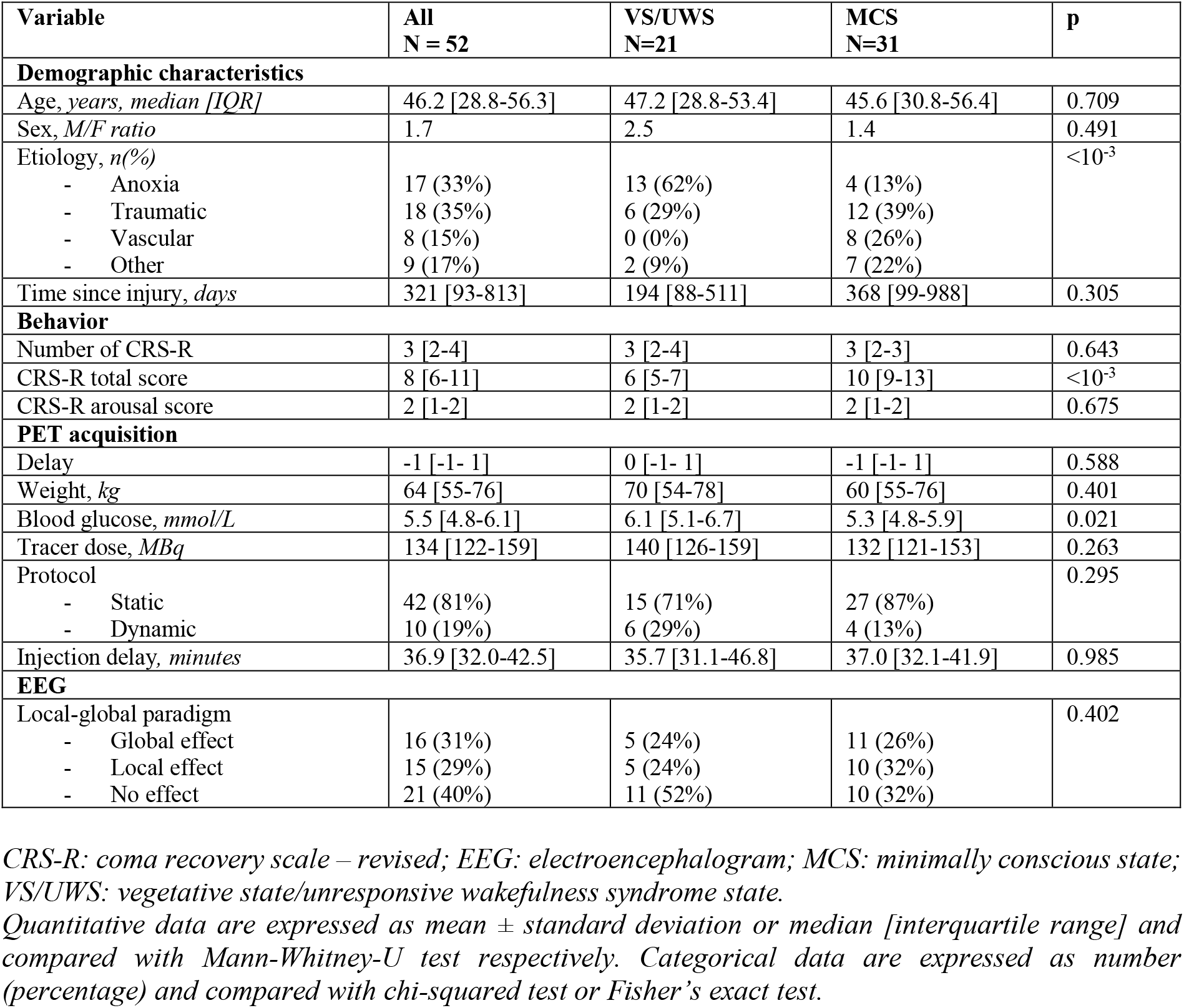
EEG/PET population characteristics

| Variable | All<br>N = 52 | VS/UWS<br>N=21 | MCS<br>N=31 | p |
| --- | --- | --- | --- | --- |
| **Demographic characteristics** |  |  |  |  |
| Age, years, median [IQR] | 46.2 [28.8-56.3] | 47.2 [28.8-53.4] | 45.6 [30.8-56.4] | 0.709 |
| Sex, M/F ratio | 1.7 | 2.5 | 1.4 | 0.491 |
| Etiology, n(%) |  |  |  | <10<sup>-3</sup> |
| - Anoxia | 17 (33%) | 13 (62%) | 4 (13%) |  |
| - Traumatic | 18 (35%) | 6 (29%) | 12 (39%) |  |
| - Vascular | 8 (15%) | 0 (0%) | 8 (26%) |  |
| - Other | 9 (17%) | 2 (9%) | 7 (22%) |  |
| Time since injury, days | 321 [93-813] | 194 [88-511] | 368 [99-988] | 0.305 |
| **Behavior** |  |  |  |  |
| Number of CRS-R | 3 [2-4] | 3 [2-4] | 3 [2-3] | 0.643 |
| CRS-R total score | 8 [6-11] | 6 [5-7] | 10 [9-13] | <10<sup>-3</sup> |
| CRS-R arousal score | 2 [1-2] | 2 [1-2] | 2 [1-2] | 0.675 |
| **PET acquisition** |  |  |  |  |
| Delay | -1 [-1- 1] | 0 [-1- 1] | -1 [-1- 1] | 0.588 |
| Weight, kg | 64 [55-76] | 70 [54-78] | 60 [55-76] | 0.401 |
| Blood glucose, mmol/L | 5.5 [4.8-6.1] | 6.1 [5.1-6.7] | 5.3 [4.8-5.9] | 0.021 |
| Tracer dose, MBq | 134 [122-159] | 140 [126-159] | 132 [121-153] | 0.263 |
| Protocol |  |  |  | 0.295 |
| - Static | 42 (81%) | 15 (71%) | 27 (87%) |  |
| - Dynamic | 10 (19%) | 6 (29%) | 4 (13%) |  |
| Injection delay, minutes | 36.9 [32.0-42.5] | 35.7 [31.1-46.8] | 37.0 [32.1-41.9] | 0.985 |
| **EEG** |  |  |  |  |
| Local-global paradigm |  |  |  | 0.402 |
| - Global effect | 16 (31%) | 5 (24%) | 11 (26%) |  |
| - Local effect | 15 (29%) | 5 (24%) | 10 (32%) |  |
| - No effect | 21 (40%) | 11 (52%) | 10 (32%) |  |

### 2. Influence of blood glucose and PET scanning protocol on FDG-PET diagnostic performances

Since blood glucose levels differed significantly between VS/UWS and MCS patients (5.9 [5.2-6.6] vs. 5.4 [4.8-6.1] respectively, p=0.019), we tested if it could have impacted the FDG-PET MIBH diagnostic performances, by including it as a covariate in a logistic regression model with the state of consciousness as the dependent variable and MIBH and blood glucose as the independent variable (state of consciousness ~ MIBH + blood glucose). We found that including blood glucose in the model resulted in very similar discrimination performances than the one obtained without blood glucose: 0.824 CI95% [0.698-0.925] vs. 0.821 [0.694-0.930], D=-0.03, p=0.976. MIBH stayed significantly associated with the clinical state of consciousness (6.2 [2.2-22.6], p=0.002) while blood glucose was not (0.7 [0.3-1.4], p=0.279).

We also checked if PET scanning protocol, that is static (single 15 minutes acquisition) or dynamic three consecutive 5 minutes acquisitions), impacted the results. The proportion of static and dynamic protocols did not differed significantly between VS/UWS and MCS groups (17/23 (74%) VS/UWS vs. 28/34 (82%) of MCS patients underwent a static scanning protocol, p=0.663). Including scanning protocol in the model did not change the model performance (0.829 CI95% [0.702-0.934] vs. 0.821 [0.694-0.930], D=-0.10, p=0.9211), with a significant association of the MIBH with the clinical state of consciousness (adjusted odds-ratio of 7.8 [2.8-28.3], p<0.001) and no association with the scanning protocol (adjusted odds-ratio 2.8 [0.5-17.0], p=0.2252).

### 3. PET regional metabolism discrimination performances (Supplementary Table 2)

**Supplementary Table 2.** PET regional metabolism discrimination performances

| LOBE | REGIONS | LEFT |  | RIGHT |  |
| --- | --- | --- | --- | --- | --- |
|  |  | AUC [95% CI] | p-fdr | AUC [95% CI] | p-fdr |
| FRONTAL | Precentral | 0.776 [0.658-0.872] | 0.0006 | 0.798 [0.675-0.899] | 0.0001 |
|  | Frontal_Sup | 0.756 [0.635-0.855] | 0.0005 | 0.783 [0.660-0.884] | 0.0004 |
|  | Frontal_Sup_Orb | 0.751 [0.630-0.851] | 0.0014 | 0.747 [0.621-0.854] | 0.0013 |
|  | Frontal_Mid | 0.729 [0.604-0.834] | 0.0041 | 0.784 [0.664-0.886] | 0.0002 |
|  | Frontal_Mid_Orb | 0.731 [0.607-0.837] | 0.0017 | 0.756 [0.634-0.863] | 0.0014 |
|  | Frontal_Inf_Oper | 0.702 [0.573-0.811] | 0.0085 | 0.780 [0.663-0.878] | 0.0002 |
|  | Frontal_Inf_Tri | 0.716 [0.590-0.821] | 0.0051 | 0.776 [0.658-0.877] | 0.0001 |
|  | Frontal_Inf_Orb | 0.730 [0.604-0.834] | 0.0022 | 0.762 [0.641-0.862] | 0.0007 |
|  | Rolandic_Oper | 0.701 [0.568-0.817] | 0.0091 | 0.762 [0.636-0.869] | 0.0004 |
| | Supp_Motor_Area | 0.817 [0.699-0.911] | $<10^{-4}$ | 0.816 [0.701-0.909] | $<10^{-4}$ |
|  | Olfactory | 0.757 [0.633-0.864] | 0.0005 | 0.758 [0.637-0.862] | 0.0009 |
|  | Frontal_Sup_Medial | 0.779 [0.656-0.881] | 0.0004 | 0.786 [0.668-0.889] | 0.0005 |
|  | Frontal_Med_Orb | 0.817 [0.701-0.909] | 0.0001 | 0.793 [0.678-0.889] | 0.0001 |
| | Rectus | 0.792 [0.677-0.889] | $<10^{-4}$ | 0.766 [0.646-0.866] | 0.0005 |
| PARIETAL | Postcentral | 0.783 [0.665-0.878] | 0.0002 | 0.795 [0.678-0.893] | $<10^{-4}$ |
|  | Parietal_Sup | 0.754 [0.635-0.853] | 0.0013 | 0.767 [0.640-0.876] | 0.0005 |
|  | Parietal_Inf | 0.682 [0.548-0.793] | 0.0179 | 0.770 [0.647-0.873] | 0.0004 |
|  | SupraMarginal | 0.662 [0.527-0.771] | 0.0375 | 0.776 [0.653-0.878] | 0.0003 |
|  | Angular | 0.664 [0.527-0.773] | 0.0347 | 0.762 [0.639-0.864] | 0.0009 |
|  | Precuneus | 0.821 [0.705-0.912] | <10<sup>-4</sup> | 0.779 [0.656-0.877] | 0.0004 |
|  | Paracentral_Lobule | 0.835 [0.730-0.919] | <10<sup>-4</sup> | 0.788 [0.666-0.887] | 0.0001 |
| TEMPORAL | Heschl | 0.720 [0.588-0.830] | 0.0044 | 0.762 [0.639-0.868] | 0.0002 |
|  | Temporal_Sup | 0.707 [0.579-0.820] | 0.0076 | 0.803 [0.688-0.900] | 0.0001 |
|  | Temporal_Pole_Sup | 0.731 [0.604-0.846] | 0.0028 | 0.772 [0.652-0.872] | 0.0005 |
|  | Temporal_Mid | 0.717 [0.588-0.821] | 0.0058 | 0.780 [0.660-0.879] | 0.0004 |
|  | Temporal_Pole_Mid | 0.696 [0.563-0.804] | 0.0093 | 0.737 [0.616-0.845] | 0.0016 |
|  | Temporal_Inf | 0.753 [0.631-0.851] | 0.0008 | 0.772 [0.648-0.873] | 0.0007 |
| OCCIPITAL | Calcarine | 0.832 [0.728-0.922] | <10<sup>-4</sup> | 0.788 [0.674-0.886] | 0.0002 |
|  | Cuneus | 0.806 [0.691-0.900] | <10<sup>-4</sup> | 0.774 [0.655-0.875] | 0.0002 |
|  | Lingual | 0.834 [0.731-0.918] | <10<sup>-4</sup> | 0.801 [0.686-0.899] | 0.0001 |
|  | Occipital_Sup | 0.765 [0.647-0.864] | 0.0004 | 0.811 [0.699-0.901] | 0.0002 |
|  | Occipital_Mid | 0.730 [0.604-0.834] | 0.0023 | 0.815 [0.704-0.903] | 0.0001 |
|  | Occipital_Inf | 0.788 [0.671-0.881] | 0.0002 | 0.772 [0.653-0.868] | 0.0006 |
|  | Fusiform | 0.751 [0.627-0.853] | 0.0012 | 0.821 [0.706-0.912] | <10<sup>-4</sup> |
| OTHER | Insula | 0.728 [0.601-0.835] | 0.0033 | 0.789 [0.664-0.892] | 0.0001 |
|  | Cingulum_Ant | 0.763 [0.638-0.864] | 0.0008 | 0.780 [0.661-0.878] | 0.0006 |
|  | Cingulum_Mid | 0.797 [0.681-0.890] | <10<sup>-4</sup> | 0.773 [0.655-0.867] | 0.0006 |
|  | Cingulum_Post | 0.786 [0.672-0.876] | 0.0003 | 0.676 [0.546-0.783] | 0.0317 |
|  | Hippocampus | 0.716 [0.588-0.824] | 0.0053 | 0.765 [0.641-0.867] | 0.0018 |
|  | ParaHippocampal | 0.751 [0.627-0.855] | 0.0012 | 0.810 [0.690-0.910] | 0.0004 |
|  | Amygdala | 0.737 [0.610-0.851] | 0.0022 | 0.811 [0.685-0.917] | <10<sup>-4</sup> |

*VS/UWS from MCS discrimination AUC with its 95% CI bootstrapped confidence interval of cortical regions of the left and right hemisphere. Significance testing against 0.5 was conducted with permutation testing with false-discovery rate correction for multiple comparisons.*
*95% CI: 95% confidence interval; AUC: area under the curve; p-fdr: false-discovery rate p-value.*

### 4. Figure e-1. Metabolic correlates of CRS-R motor and arousal MCS items with liberal statistical threshold

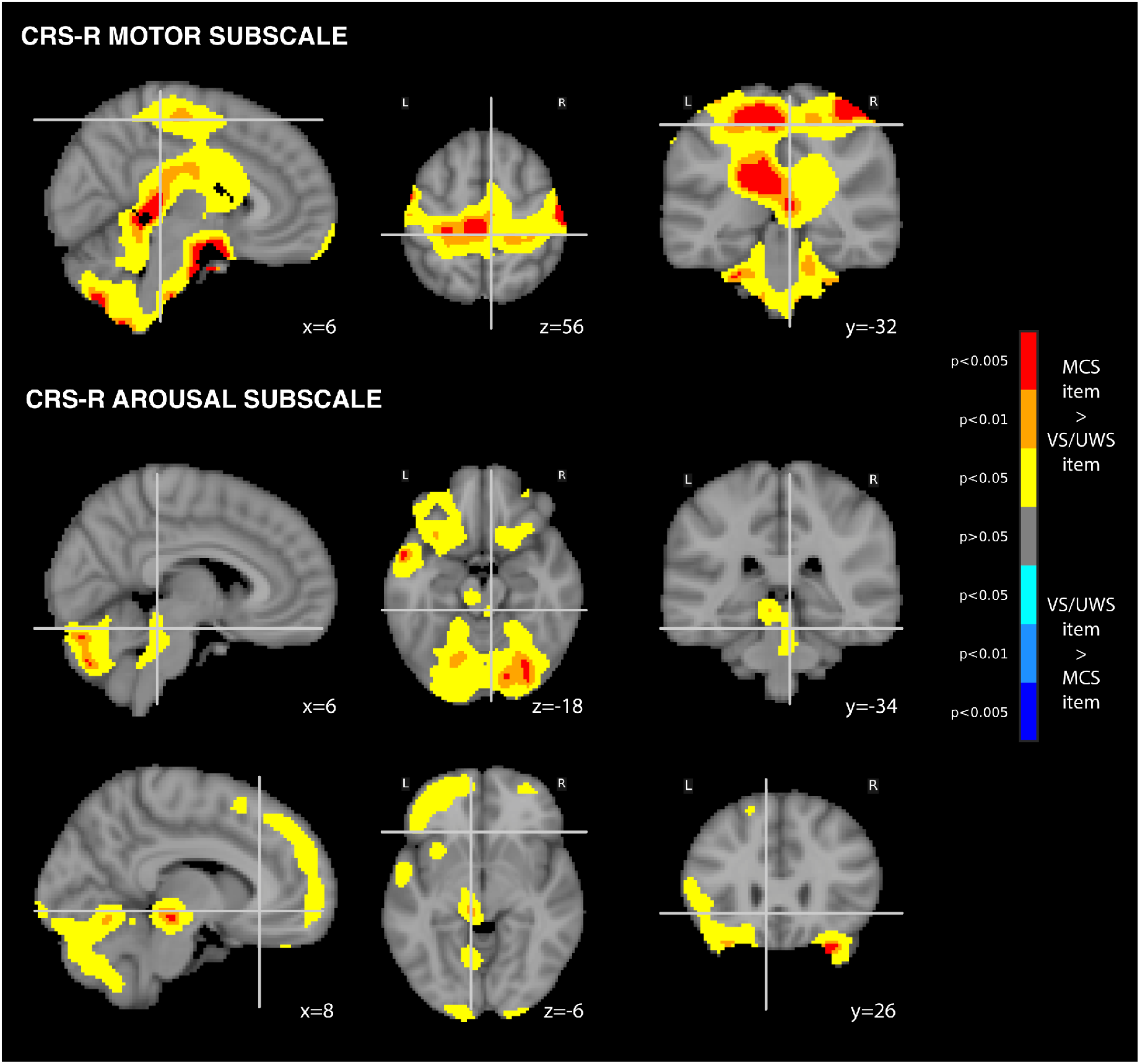
Independent FDG-PET metabolic correlates of the CRS-R MCS items in the motor subscale. **(A)** and arousal subscale **(B)** (p<0.05 uncorrected, cluster extent 100 voxels, superimposed on coronal, sagittal and axial slices of the MNI 152 T1 brain template with related y, x and z MNI coordinates). L=left; R=right.

### 5. Figure e-2. Whole-brain metabolic correlates of conscious state

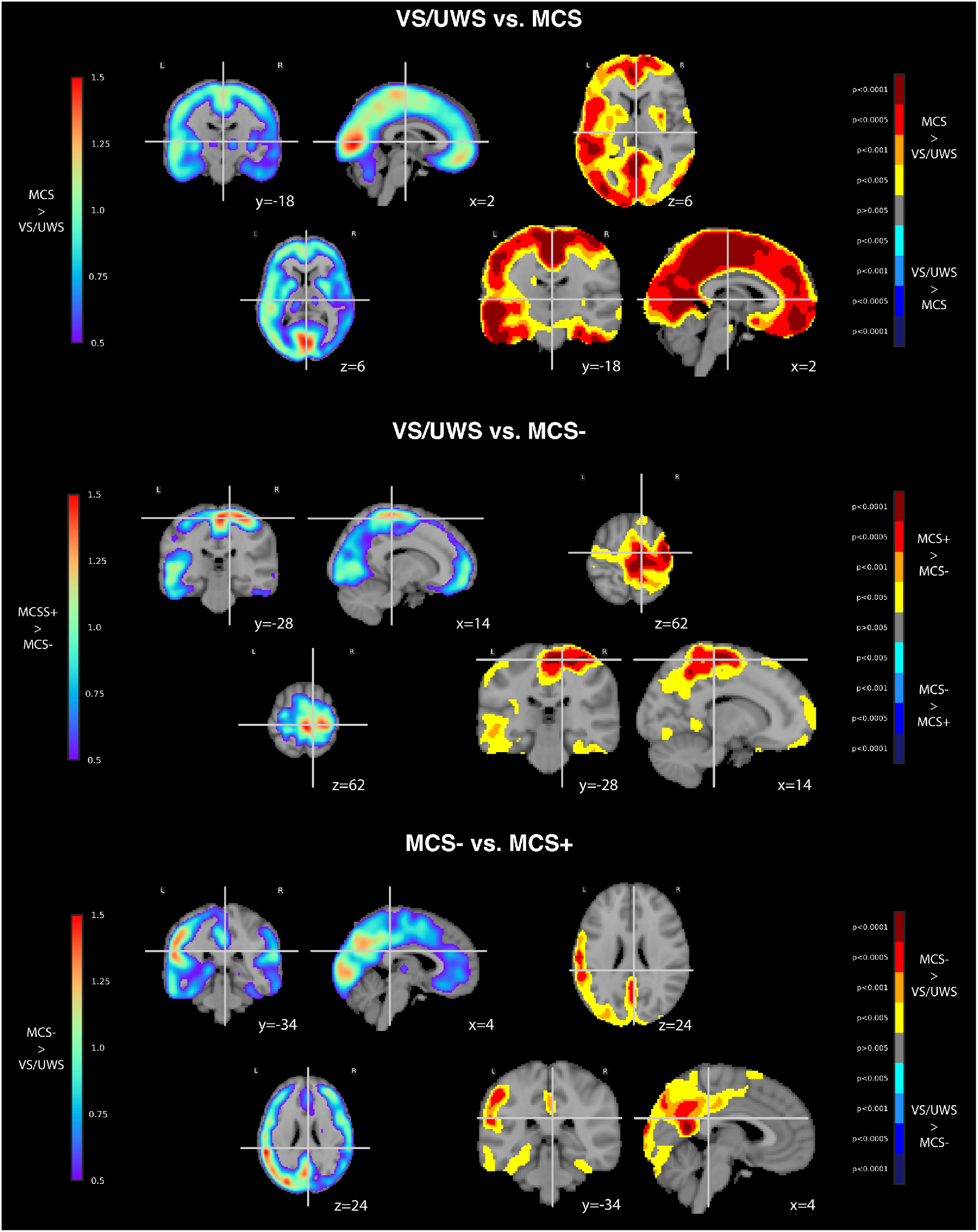
FDG-PET metabolic contrast of different state of consciousness: vegetative state/unresponsive wakefulness syndrome (VS/UWS) vs. minimally conscious state (MCS) **(A)**VS/UWS vs. MCS- **(B)** and MCS- vs. MCS+ **(C)** (p<0.005 uncorrected, cluster extent 100 voxels, superimposed on coronal, sagittal and axial slices of the MNI 152 T1 brain template with related y, x and z MNI coordinates). L=left; R=right.

## Notes

### Competing Interest Statement

The authors have declared no competing interest.

### Clinical Trial

This research is a substudy of a observational prospective protocol collecting routine care data of patients suffering from disorders of consciousness (NEURODOC protocol ethic committee approval 2013-A01385-40).

### Funding Statement

BH was funded by a "Bourse poste d'Accueil" from "Institut National de la Recherche Medicale". This work was also supported by the FRM 2015 (LN) the Academie des Sciences-Lamonica Prize 2016 (LN) and by the "Recovery of consciousness after severe brain injury Phase II" grant of the James S. McDonnell Foundation. The research leading to these results has received funding from the program "Investissements d'avenir" ANR-10-IAIHU-06.

### Author Declarations

The protocol was approved by the local ethic committee (Comite de Protection des Personnes 2013-A01385-40) Ile de France 1 (Paris France) under the code 'Recherche en soins courants' (routine care research).

